# Prevalence and Clinical Predictors of Stroke Mimics in the Extended Thrombectomy Window: A Cross-Sectional Study in Vietnam

**DOI:** 10.1101/2025.09.15.25335749

**Authors:** Tran Ba Lan, Ngo Cam Hoa, Luong Ngoc Quyen, Le Thi My Duyen

## Abstract

**Background:** Stroke mimics pose diagnostic challenges in the emergency department, particularly in patients presenting within the 4.5-24 hour thrombectomy window. Misdiagnosis may result in unnecessary interventions or delayed stroke treatment. This study aimed to determine the prevalence, clinical characteristics, and predictors of stroke mimics among suspected stroke patients in this timeframe.

**Methods:** We conducted a cross-sectional study at People’s Hospital 115, Vietnam (February-June 2024). Patients presenting with acute neurological deficits underwent standardized magnetic resonance imaging. Logistic regression identified independent predictors of stroke mimics.

**Results:** Among 1,180 patients (mean age 62.6 years; 58.6% male), 346 (29.3%) were diagnosed with SMs. Compared to stroke patients, stroke mimics were more likely to be male (77.2% vs. 50.8%, P < 0.001), had lower NIHSS scores (median 8 vs. 12.5, P = 0.013), and shorter hospital stays (median 2 vs. 4 days, P < 0.001). Independent predictors included bilateral leg weakness (OR 23.9; 95% CI, 6.9-33.8), quadriparesis (OR 25.4; 95% CI, 7.8-35.1), dizziness (OR 14.6; 95% CI, 7.1-32.7), headache (OR 13.2; 95% CI, 6.5-26.8), seizures (OR 13.9; 95% CI, 6.7-28.5), altered consciousness (OR 4.9; 95% CI, 2.4-9.8), numbness (OR 6.4; 95% CI, 2.1-12.5), and male sex (OR 3.3; 95% CI, 2.5-4.4). Atrial fibrillation was a negative predictor (OR 0.33).

**Conclusions:** Stroke mimics account for nearly one-third of suspected strokes within the extended thrombectomy window. Recognizing specific clinical predictors can improve diagnostic accuracy, reduce unnecessary interventions, and optimize ED resource allocation. Integration of these predictors into triage protocols could support faster, safer decision-making in clinical practice.

**Key Points:** *What is already known on this topic:* Stroke mimics are common and complicate triage in the era of thrombectomy; prevalence varies widely but data from Southeast Asia are scarce.

*What this study adds:* In this large Vietnamese cohort, nearly one-third were stroke mimics; distinct clinical predictors were identified.

*How this study might affect research, practice or policy:* Incorporating these predictors into triage could reduce unnecessary interventions and optimize ED resources.

## Introduction

Advances in neuroimaging and reperfusion therapies have expanded the therapeutic window for mechanical thrombectomy up to 24 hours in selected patients with large vessel occlusion (1). While this has improved outcomes, it has also increased the workload in emergency departments (EDs) (2). A proportion of patients with stroke-like symptoms, termed stroke mimics (SMs) - complicates triage and management.

SMs are defined as non-stroke conditions presenting with acute focal neurological deficits. Reported prevalence varies from 2% to 38%, depending on clinical setting, diagnostic criteria, and imaging modalities (3-5). Misdiagnosis can lead to unnecessary interventions, delays in care, and resource misallocation. MRI, with its superior sensitivity for non-stroke pathologies, tends to yield higher SM rates compared to CT.

Data on SMs in low- and middle-income countries, particularly in Southeast Asia, are limited. This study aimed to evaluate the prevalence, clinical features, and independent predictors of SMs among patients presenting within the 4.5–24-hour window in a tertiary stroke referral center in Vietnam.

## Methods

### Study Design

We performed a cross-sectional study at People’s Hospital 115, Ho Chi Minh City, from February to June 2024. This tertiary hospital manages a high volume of acute stroke cases annually.

### Participants

Inclusion criteria were adults (≥18 years) presenting with acute focal neurological deficits 4.5–24 hours after symptom onset or last known well. Exclusion criteria included prior imaging at another hospital, contraindications to MRI, or incomplete data.

### Diagnostic Workup

ED physicians activated “code stroke” for eligible patients. All underwent standardized MRI brain protocol, including axial FLAIR, T2, DWI, T1 sagittal, coronal T2-TSE, and MRA sequences (1.5T GE SIGNA Explorer). Images were reviewed by a senior neuroradiologist. Stroke mimics were defined as patients presenting with acute focal neurological deficits but without evidence of acute ischemic lesions on brain MRI. The final diagnosis of stroke mimic was established at hospital discharge based on MRI findings and consensus of treating neurologists (Figure 1).

**Figure 1.**
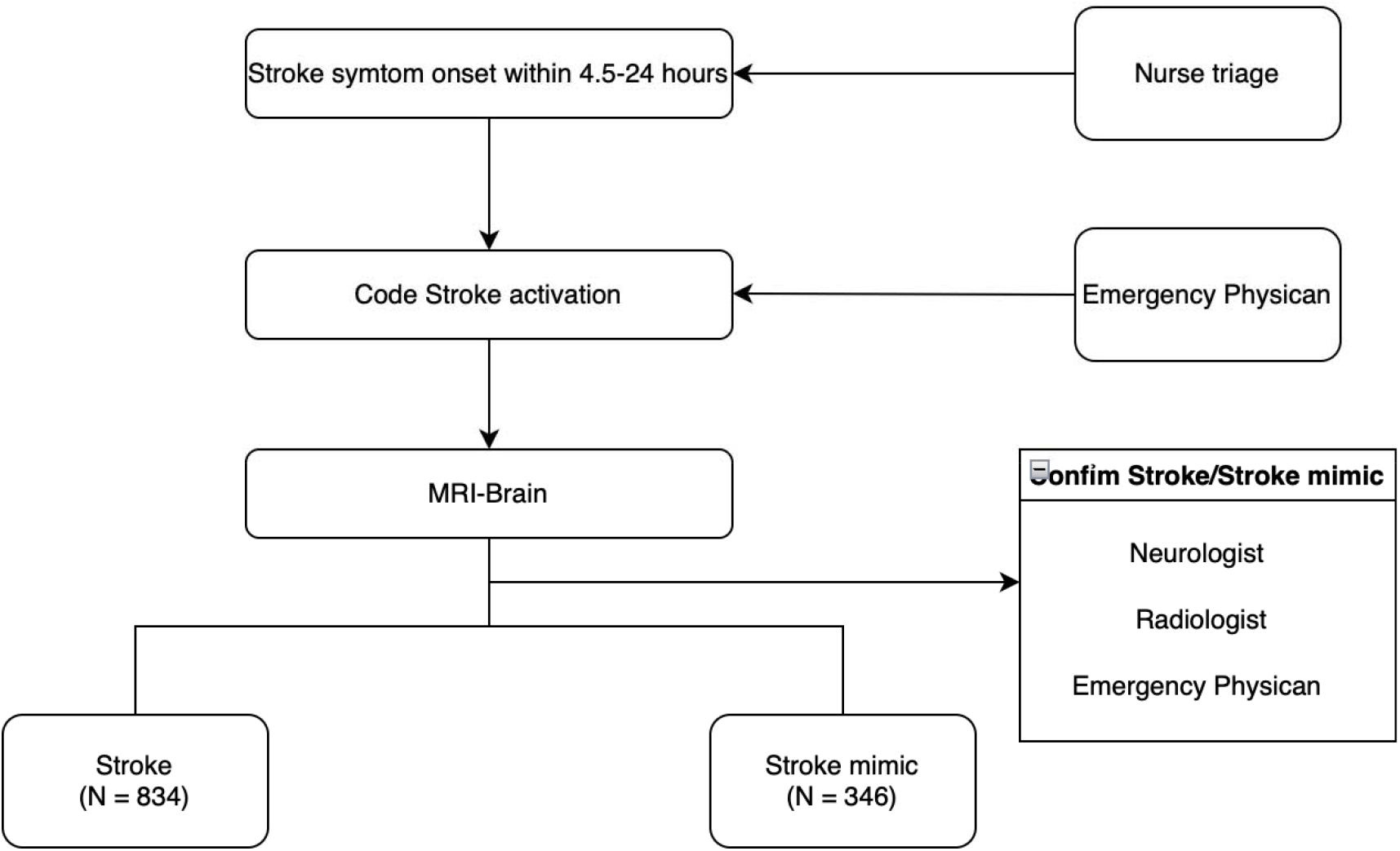
Flowchart of patient enrollment and classification into stroke versus stroke mimics. Flow diagram showing the selection of patients presenting with acute focal neurological deficits within 4.5–24 hours of onset. Patients were classified into ischemic stroke or stroke mimic groups based on MRI findings. ED: Emergency Department; MRI: Magnetic Resonance Imaging.

### Data Collection

Data included demographics, comorbidities, NIHSS scores, presenting symptoms, hospital stay, imaging results, and discharge diagnoses.

### Statistical Analyses

Continuous variables were presented as mean ± standard deviation (SD) or median [interquartile range (IQR)], and categorical variables as numbers and percentages. Between-group comparisons were performed using Student’s t test or Mann–Whitney U test for continuous variables and Chi-square or Fisher’s exact test for categorical variables. Multivariate logistic regression was conducted to identify independent predictors of stroke mimics, including variables with P < 0.10 in univariate analysis.

Missing data were rare (<5% for all variables). Analyses were performed on available cases (available-case analysis), whereby patients were excluded only from analyses requiring the missing variable. Given the very low proportion of missingness, multiple imputation was not applied, as exclusion of these cases did not materially alter the results. Analyses were performed using R (version 4.3.0).

### Ethical Considerations

The study was approved by the Institutional Review Board of People’s Hospital 115 (No. 2283). Written informed consent was obtained from all participants.

## Results

### Baseline Characteristics

Among 1,374 patients screened, 194 were excluded because MRI was not performed or data were incomplete. A total of 1,180 patiens were included in the final analysis, comprising 834 strokes and 346 SMs (Figure 1). The mean age did not differ significantly between groups (P = 0.750). However, SM patients were more frequently male (77.2% vs. 50.8% in stroke, P < 0.001) and had lower NIHSS scores at admission (median 8 vs. 12.5, P = 0.013) (Table 1). Hospital length of stay was shorter in the SM group (median 2 vs. 4 days, P < 0.001), with correspondingly lower treatment costs (P < 0.001) (Table 2).

**Table 1.** Baseline characteristics of patients with stroke mimics versus stroke.

| Characteristics | Mimics (N=346) | Strokes (N=834) | P-value |
| --- | --- | --- | --- |
| Age (years), mean ± SD | 62.71 ± 14.04 | 62.42 ± 13.53 | 0.750 |
| Male sex, n (%) | 267 (77.2%) | 424 (50.8%) | <0.001 |
| Time from arrival to MRI (min), mean ± SD | 136.76 ± 76.84 | 145.54 ± 78.71 | 0.080 |
| Length of stay in ED (min), mean ± SD | 249.50 ± 113.85 | 260.77 ± 115.67 | 0.124 |
| Time from onset to ED (min), median [IQR] | 360 [180–780] | 360 [210–690] | 0.480 |
| NIHSS score at admission, median [range] | 8 [1–25] | 12.5 [1–25] | 0.013 |
| Elevated blood pressure ≥150/90 mmHg, n (%) | 131 (37.9%) | 334 (40.1%) | 0.062 |
| Health insurance, n (%) | 314 (90.8%) | 721 (86.5%) | 0.051 |

*Values are presented as mean ± SD, median [IQR], or n (%) as appropriate. ED: Emergency Department; MRI: Magnetic Resonance Imaging; NIHSS: National Institutes of Health Stroke Scale.*

**Table 2.**
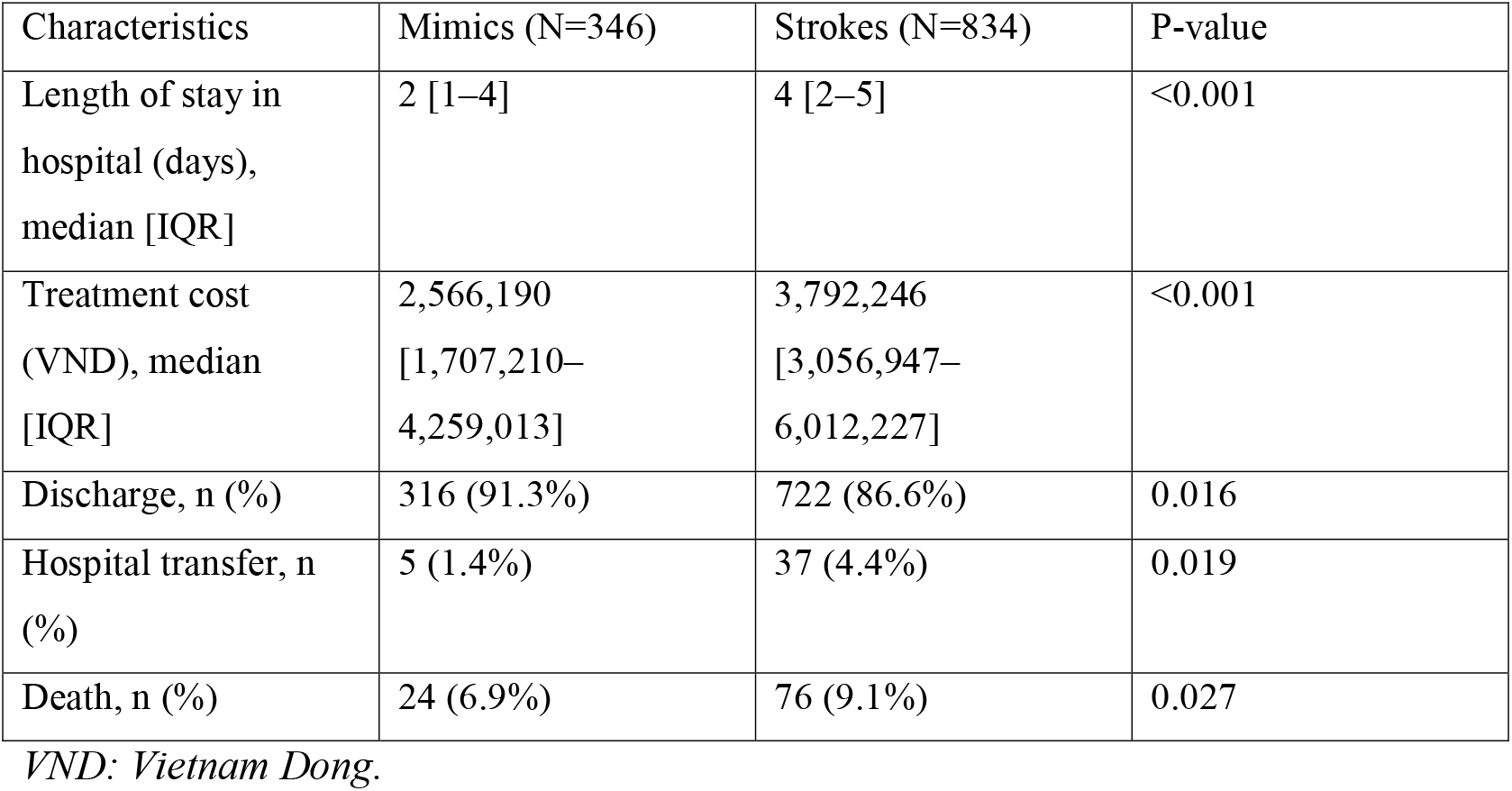
In-hospital outcomes and healthcare utilization of patients with stroke mimics versus stroke.

| Characteristics | Mimics (N=346) | Strokes (N=834) | P-value |
| --- | --- | --- | --- |
| Length of stay in hospital (days), median [IQR] | 2 [1–4] | 4 [2–5] | <0.001 |
| Treatment cost (VND), median [IQR] | 2,566,190<br>[1,707,210–<br>4,259,013] | 3,792,246<br>[3,056,947–<br>6,012,227] | <0.001 |
| Discharge, n (%) | 316 (91.3%) | 722 (86.6%) | 0.016 |
| Hospital transfer, n (%) | 5 (1.4%) | 37 (4.4%) | 0.019 |
| Death, n (%) | 24 (6.9%) | 76 (9.1%) | 0.027 |

*VND: Vietnam Dong.*

### Clinical Presentation

Compared with stroke, SM patients more often presented with atypical symptoms, including headache (7.5% vs. 1.3%, P < 0.001), dizziness (9.5% vs. 1.2%, P < 0.001), seizures (7.2% vs.

1.0%, P < 0.001), and altered consciousness (28.0% vs. 10.3%, P < 0.001). Bilateral leg weakness (2.6% vs. 0.4%, P = 0.003), quadriparesis (6.1% vs. 0.7%, P < 0.001), and numbness (1.4% vs. 0.4%, P = 0.011) were also significantly more common among SM patients (Table S1).

### Etiologies of Stroke Mimics

The leading causes of SMs were sepsis (13%), peripheral vertigo (11%), post-stroke sequelae (10%), and radiculopathy (10%). Other etiologies included epilepsy (9%), Guillain–Barré syndrome (7%), headache disorders (7%), and electrolyte disturbances (5%). Functional neurological disorders, hepatic encephalopathy, and miscellaneous conditions accounted for the remainder (Table 3).

**Table 3.**
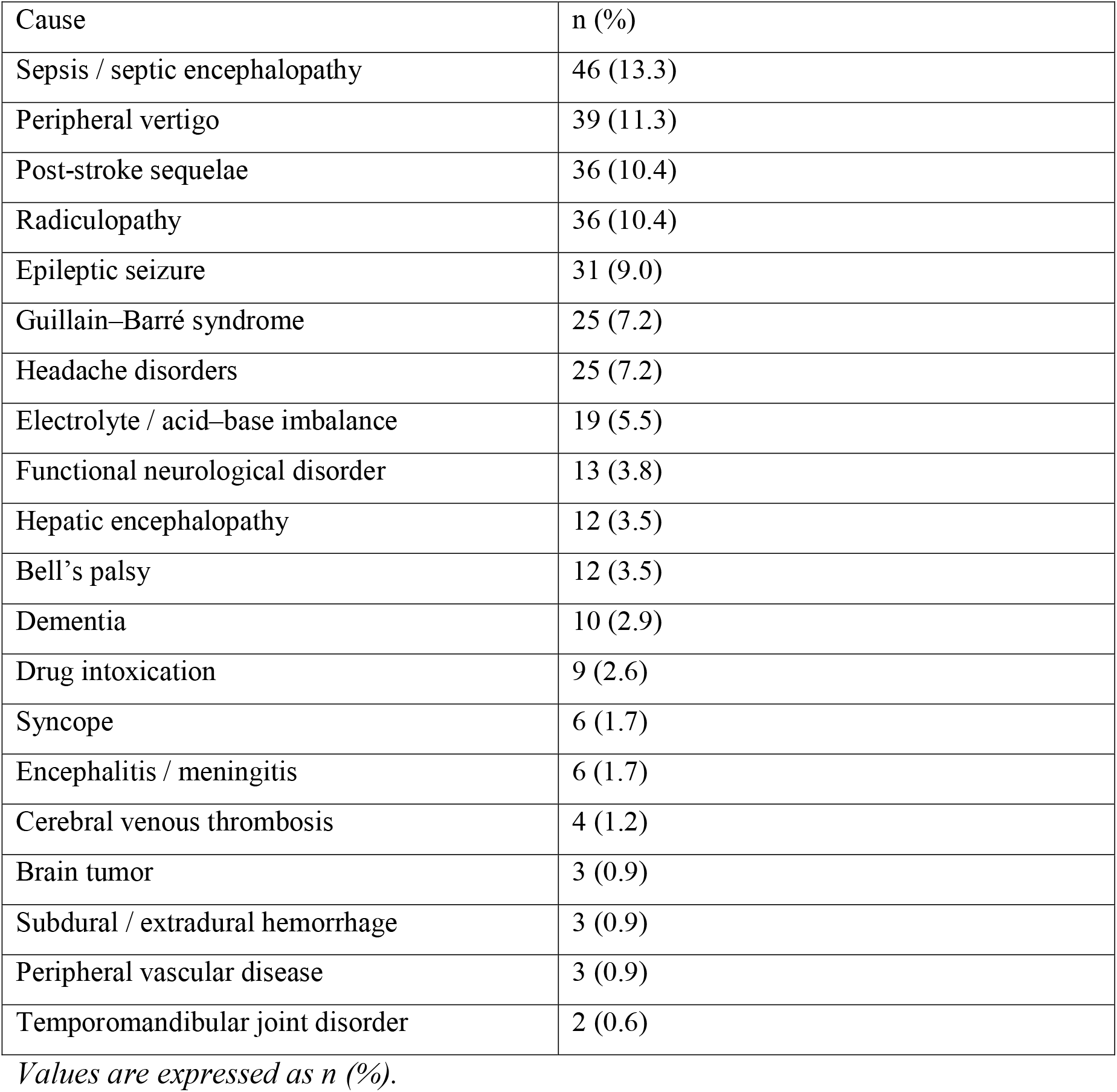
Etiologies of stroke mimics (n = 346)

| Cause | n (%) |
| --- | --- |
| Sepsis / septic encephalopathy | 46 (13.3) |
| Peripheral vertigo | 39 (11.3) |
| Post-stroke sequelae | 36 (10.4) |
| Radiculopathy | 36 (10.4) |
| Epileptic seizure | 31 (9.0) |
| Guillain–Barré syndrome | 25 (7.2) |
| Headache disorders | 25 (7.2) |
| Electrolyte / acid–base imbalance | 19 (5.5) |
| Functional neurological disorder | 13 (3.8) |
| Hepatic encephalopathy | 12 (3.5) |
| Bell’s palsy | 12 (3.5) |
| Dementia | 10 (2.9) |
| Drug intoxication | 9 (2.6) |
| Syncope | 6 (1.7) |
| Encephalitis / meningitis | 6 (1.7) |
| Cerebral venous thrombosis | 4 (1.2) |
| Brain tumor | 3 (0.9) |
| Subdural / extradural hemorrhage | 3 (0.9) |
| Peripheral vascular disease | 3 (0.9) |
| Temporomandibular joint disorder | 2 (0.6) |

*Values are expressed as n (%).*

### Predictors of Stroke Mimics

Univariate analysis demonstrated significant associations between SMs and male sex, headache, dizziness, seizures, bilateral leg weakness, quadriparesis, numbness, and altered consciousness (all P < 0.01) (Table S3). In multivariate logistic regression, independent predictors included quadriparesis (OR 25.41; 95% CI, 7.8–35.1), bilateral leg weakness (OR 23.86; 95% CI, 6.9– 33.8), dizziness (OR 14.62; 95% CI, 7.1–32.7), seizures (OR 13.90; 95% CI, 6.7–28.5), headache (OR 13.15; 95% CI, 6.5–26.8), numbness (OR 6.38; 95% CI, 2.1–12.5), altered consciousness (OR 4.85; 95% CI, 2.4–9.8), and male sex (OR 3.27; 95% CI, 2.47–4.37). Atrial fibrillation emerged as a negative predictor (OR 0.33; 95% CI, 0.15–0.68) (Table 4).

**Table 4.**
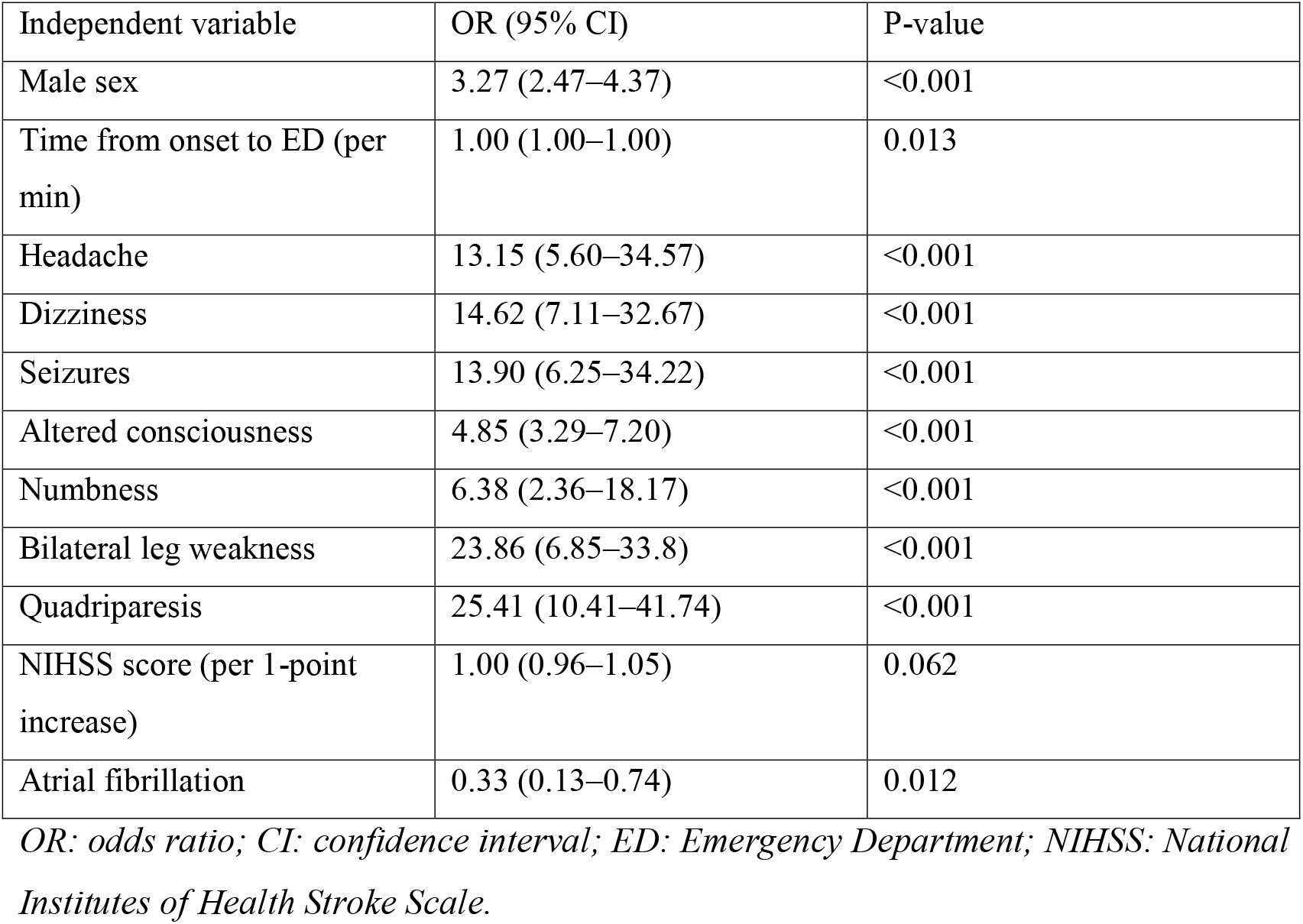
Multivariate logistic regression analysis for independent predictors of stroke mimics.

## Discussion

This cross-sectional study of 1,180 patients presenting within the extended thrombectomy window found that nearly one-third (29.3%) were diagnosed with SMs. Compared with strokes, mimics were more often male, had lower NIHSS scores, and presented with atypical symptoms such as headache, dizziness, seizure, or altered consciousness. Stroke mimic patients also had shorter hospital stays, lower treatment costs, and lower in-hospital mortality. Multivariate analysis identified headache, seizure, altered consciousness, and bilateral weakness as independent predictors of stroke mimics.

### Prevalence and Characteristic of Stroke Mimics

In our cohort, SMs accounted for nearly one-third of patients presenting within the 4.5–24-hour window, a prevalence consistent with the upper range reported in prior studies (2–38%) (6-8). Variability in SM prevalence is influenced by several factors, including the evaluating clinician (7), the definition of stroke mimics (9-12), and the choice of initial imaging modality. Studies relying on MRI for diagnostic tend to report higher SM rates compared to those utilizing CT or CT angiography (3, 13-16).

The higher mimic rates in MRI-based studies demonstrate its superior ability to detect non-stroke conditions, such as brain tumours, encephalitis, and metabolic encephalopathies, which can be missed by CT (17). This highlights MRI’s value in distinguishing strokes from mimics, especially in cases with atypical presentations or early ischemic changes invisible on CT.

### Etiologies of Stroke Mimics

The etiologies of SMs in our study were heterogeneous, with sepsis emerging as the leading cause (13%), followed by peripheral vertigo, post-stroke sequelae, and radiculopathy. This contrasts with Western series where functional disorders, migraine, or vestibular syndromes predominate (3, 9, 16).

The prominence of sepsis in our population likely reflects both the tertiary referral nature of our center and the high prevalence of systemic infections in critically ill patients. Altered consciousness – common in our SM cohort was frequently linked to septic presentations, underscoring the importance of considering systemic conditions when evaluating ambiguous neurological deficits.

Outcomes for SM patients were more favorable than for stroke patients, with shorter hospitalizations, lower costs, and higher discharge rates. These results emphasize the importance of accurate differentiation, which can minimize unnecessary interventions and optimize ED resource utilization.

### Predictors of Stroke mimics

Our regression analysis identified nine independent predictors of SMs, including quadriparesis, bilateral leg weakness, dizziness, seizures, headache, numbness, altered consciousness, and male sex, while atrial fibrillation was a negative predictor. These findings are partially aligned with prior literature, where predictors have included low NIHSS, younger age, female sex, and decreased consciousness (4, 13, 16, 18-20).

The association with male sex is noteworthy. In our study, 77.2% of SMs were male, contrasting with previous reports where SMs were more common in women, often due to functional disorders (21, 22). In our cohort, functional neurological disorders accounted for only 3.8% of SM cases, whereas sepsis and septic shock with higher incidence in men were the leading etiologies. This demographic pattern likely explains the observed sex difference and highlights the role of population-specific factors (23, 24). The strong negative association with atrial fibrillation (OR 0.33) reinforces its close link with true ischemic stroke and may aid clinicians in rapid risk stratification.

### Clinical Implications

Our findings suggest that clinicians should maintain heightened suspicion for SMs when patients present with atypical features such as bilateral leg weakness, quadriparesis, or dizziness. Early MRI in such cases may improve diagnostic accuracy, reduce unnecessary interventions, and streamline ED workflows. Integrating clinical predictors with imaging algorithms could enhance triage decisions in extended thrombectomy windows.

In summary, nearly one-third of patients presenting within the extended thrombectomy window were diagnosed with SMs. These patients more often had atypical symptoms, better short-term outcomes, and lower mortality than ischemic strokes. Independent predictors included headache, seizure, altered consciousness, and bilateral weakness. While informative, these results should be interpreted with caution given the single-center design and exclusion of patients without MRI. The generalisability of these findings is limited by the single-center design. However, the large sample size and standardized MRI-based assessment support their applicability to similar emergency and stroke centers in Vietnam and comparable healthcare systems.

### Limitations

This study has several limitations. First, it was conducted at a single tertiary center, which may restrict the generalisability of our findings to other healthcare settings. Second, the diagnostic process might have missed patients with transient ischemic attacks or very small ischemic lesions, which can be difficult to detect even on MRI. Third, patients who did not undergo MRI were excluded, potentially introducing selection bias and underestimating the true prevalence of SMs. Finally, we only assessed in-hospital outcomes; long-term prognosis of SMs could not be determined within this study design.

## CONCLUSION

Stroke mimics accounted for nearly one-third of suspected strokes in the extended thrombectomy window. Recognizing key clinical predictors may improve diagnostic accuracy and optimize emergency care. Further multicenter studies are needed to validate these findings.

## Supporting information

Table S1. Clinical presentation of stroke mimics versus stroke

Table S2. Vascular risk factors of patients with stroke mimics versus stroke

Table S3. Univariate analysis of factors associated with stroke mimics

## Data Availability

All data produced in the present study are available upon reasonable request to the corresponding author.

## ACKNOWLEDGMENTS

We thank all study participants and the staff of People’s Hospital 115 for their assistance in data collection and analysis.

## PATIENT AND PUBLIC INVOLVEMENT

Patients and the public were not involved in the design, conduct, reporting, or dissemination plans of this research.

## REFERENCES

1. Kleindorfer DO, Towfighi A, Chaturvedi S, Cockroft KM, Gutierrez J, Lombardi-Hill D, et al. 2021 Guideline for the Prevention of Stroke in Patients With Stroke and Transient Ischemic Attack: A Guideline From the American Heart Association/American Stroke Association. Stroke. 2021;52(7):e364–e467.

2. Warner JJ, Harrington RA, Sacco RL, Elkind MSV. Guidelines for the Early Management of Patients With Acute Ischemic Stroke: 2019 Update to the 2018 Guidelines for the Early Management of Acute Ischemic Stroke. Stroke. 2019;50(12):3331–2.

3. Quenardelle V, Lauer-Ober V, Zinchenko I, Bataillard M, Rouyer O, Beaujeux R, et al. Stroke Mimics in a Stroke Care Pathway Based on MRI Screening. Cerebrovasc Dis. 2016;42(3-4):205–12.

4. Hand PJ, Kwan J, Lindley RI, Dennis MS, Wardlaw JM. Distinguishing between stroke and mimic at the bedside: the brain attack study. Stroke. 2006;37(3):769–75.

5. Merino JG, Luby M, Benson RT, Davis LA, Hsia AW, Latour LL, et al. Predictors of acute stroke mimics in 8187 patients referred to a stroke service. J Stroke Cerebrovasc Dis. 2013;22(8):e397–403.

6. Zinkstok SM, Engelter ST, Gensicke H, Lyrer PA, Ringleb PA, Artto V, et al. Safety of thrombolysis in stroke mimics: results from a multicenter cohort study. Stroke. 2013;44(4):1080–4.

7. Sjoo M, Berglund A, Sjostrand C, Eriksson EE, Mazya MV. Prehospital stroke mimics in the Stockholm Stroke Triage System. Front Neurol. 2022;13:939618.

8. Kim T, Jeong HY, Suh GJ. Clinical Differences Between Stroke and Stroke Mimics in Code Stroke Patients. J Korean Med Sci. 2022;37(7):e54.

9. Al Khathaami AM, Alsaif SA, Al Bdah BA, Alhasson MA, Aldriweesh MA, Alluhidan WA, et al. Stroke mimics: Clinical characteristics and outcome. Neurosciences (Riyadh). 2020;25(1):38–42.

10. Nguyen PL, Chang JJ. Stroke Mimics and Acute Stroke Evaluation: Clinical Differentiation and Complications after Intravenous Tissue Plasminogen Activator. J Emerg Med. 2015;49(2):244–52.

11. Long B, Koyfman A. Clinical Mimics: An Emergency Medicine-Focused Review of Stroke Mimics. J Emerg Med. 2017;52(2):176–83.

12. Chtaou N, Bouchal S, Midaoui AEL, Souirti Z, Tachfouti N, Belahsen MF. Stroke Mimics: Experience of a Moroccan Stroke Unit. J Stroke Cerebrovasc Dis. 2020;29(5):104651.

13. Ifergan H, Amelot A, Ismail M, Gaudron M, Cottier JP, Narata AP. Stroke-mimics in stroke-units. Evaluation after changes imposed by randomized trials. Arq Neuropsiquiatr. 2020;78(2):88–95.

14. Cramer SC, Stradling D, Brown DM, Carrillo-Nunez IM, Ciabarra A, Cummings M, et al. Organization of a United States county system for comprehensive acute stroke care. Stroke. 2012;43(4):1089–93.

15. Prodi E, Danieli L, Manno C, Pagnamenta A, Pravata E, Roccatagliata L, et al. Stroke Mimics in the Acute Setting: Role of Multimodal CT Protocol. AJNR Am J Neuroradiol. 2022;43(2):216–22.

16. Pohl M, Hesszenberger D, Kapus K, Meszaros J, Feher A, Varadi I, et al. Ischemic stroke mimics: A comprehensive review. J Clin Neurosci. 2021;93:174–82.

17. Boulter DJ, Schaefer PW. Stroke and stroke mimics: a pattern-based approach. Semin Roentgenol. 2014;49(1):22–38.

18. Libman RB, Wirkowski E, Alvir J, Rao TH. Conditions that mimic stroke in the emergency department. Implications for acute stroke trials. Arch Neurol. 1995;52(11):1119–22.

19. Okano Y, Ishimatsu K, Kato Y, Yamaga J, Kuwahara K, Okumoto K, et al. Clinical features of stroke mimics in the emergency department. Acute Med Surg. 2018;5(3):241–8.

20. Jones AT, O’Connell NK, David AS. Epidemiology of functional stroke mimic patients: a systematic review and meta-analysis. Eur J Neurol. 2020;27(1):18–26.

21. Hallett M, Aybek S, Dworetzky BA, McWhirter L, Staab JP, Stone J. Functional neurological disorder: new subtypes and shared mechanisms. Lancet Neurol. 2022;21(6):537–50.

22. Baizabal-Carvallo JF, Jankovic J. Gender Differences in Functional Movement Disorders. Mov Disord Clin Pract. 2020;7(2):182–7.

23. Esper AM, Moss M, Lewis CA, Nisbet R, Mannino DM, Martin GS. The role of infection and comorbidity: Factors that influence disparities in sepsis. Crit Care Med. 2006;34(10):2576–82.

24. Martin GS, Mannino DM, Eaton S, Moss M. The epidemiology of sepsis in the United States from 1979 through 2000. N Engl J Med. 2003;348(16):1546–54.

