## Supplementary material for "Prevalence and Clinical Predictors of Stroke Mimics in the Extended Thrombectomy Window: A Cross-Sectional Study in Vietnam": Table S1. Clinical presentation of stroke mimics versus stroke

| Characteristics | Mimics (N=346) | Strokes (N=834) | P-value |
| --- | --- | --- | --- |
| Headache, n (%) | 26 (7.5%) | 11 (1.3%) | <0.001 |
| Dizziness, n (%) | 33 (9.5%) | 10 (1.2%) | <0.001 |
| Seizures, n (%) | 25 (7.2%) | 8 (1.0%) | <0.001 |
| Altered consciousness, n (%) | 97 (28.0%) | 86 (10.3%) | <0.001 |
| Numbness, n (%) | 5 (1.4%) | 3 (0.4%) | 0.011 |
| Bilateral leg weakness, n (%) | 9 (2.6%) | 3 (0.4%) | 0.003 |
| Quadriparesis, n (%) | 21 (6.1%) | 6 (0.7%) | <0.001 |

*Values are presented as n (%.*

*ED: Emergency Department; NIHSS: National Institutes of Health Stroke Scale.*
