## Supplementary material for "Prevalence and Clinical Predictors of Stroke Mimics in the Extended Thrombectomy Window: A Cross-Sectional Study in Vietnam": Table S2. Vascular risk factors of patients with stroke mimics versus stroke

| Characteristics | Mimics (N=346) | Strokes (N=834) | P-value |
| --- | --- | --- | --- |
| Hypertension, n (%) | 248 (71.7%) | 586 (70.3%) | 0.678 |
| Diabetes mellitus, n (%) | 81 (23.4%) | 204 (24.5%) | 0.757 |
| Dyslipidemia, n (%) | 126 (36.4%) | 321 (38.5%) | 0.547 |
| Atrial fibrillation, n (%) | 7 (2.0%) | 44 (5.3%) | 0.019 |

*Values are presented as n (%).*
