## Supplementary material for "Prevalence and Clinical Predictors of Stroke Mimics in the Extended Thrombectomy Window: A Cross-Sectional Study in Vietnam": Table S3. Univariate analysis of factors associated with stroke mimics

| Variable | P-value |
| --- | --- |
| Age | 0.748 |
| Male sex | <0.001 |
| Headache | <0.001 |
| Dizziness | <0.001 |
| Seizures | <0.001 |
| Altered consciousness | <0.001 |
| Fever | 0.984 |
| Numbness | 0.011 |
| Bilateral leg weakness | 0.003 |
| Quadriparesis | <0.001 |
| NIHSS score | 0.013 |
| Time from onset to ED | 0.002 |
| Hypertension | 0.063 |
| Diabetes mellitus | 0.701 |
| Dyslipidemia | 0.504 |
| Ischemic heart disease | 0.355 |
| Sequelae of cerebrovascular disease | 0.615 |
| Peripheral vascular disease | 0.965 |
| Atrial fibrillation | 0.002 |

*Variables with P < 0.05 were considered statistically significant.*
